# Exploring factors for meaningful patient involvement in infectious disease clinical studies: A qualitative pilot study among key stakeholders

**DOI:** 10.1101/2023.09.01.23294944

**Authors:** S. Moggré, T. ten Doesschate, E. Sieswerda, K.A.G.J. Romijnders

## Abstract

**Introduction:** The attitude towards patient involvement in clinical research has changed dramatically over the years, from research about patient to research with patients. This qualitative study aimed to explore perceptions, ideas, and opinions of stakeholders about integrating the patient perspective into clinical research in infectious diseases in the Netherlands.

**Methods:** Stakeholders involved with clinical research in infectious diseases were purposefully sampled between March and June 2023. Interviews were conducted using a semi-structured guide based on the Consolidated Framework for Intervention Research and feasibility framework.

**Results:** Of the thirteen stakeholders, six were (clinical) researchers, two represented pharmaceutical companies, two were involved with policy making, and three were patient representatives. Patient involvement in the design and conduct of clinical research in infectious diseases was seen as crucial, although the mode of involvement could differ between research in acute and chronic infections. Stakeholders observed a gap among patients and clinical researchers, which was believed to lead to a phenomenon described as an ivory tower. Key opinion leaders may potentially bridge these barriers and serve as protagonists for meaningful patient involvement. Stakeholders acknowledged the need of communication and expertise to integrate the patient perspective in clinical research in infectious diseases.

**Conclusion:** Our qualitative analysis underlines that despite barriers, such as communication and expertise, stakeholders recognize the importance of integrating the patient perspective in clinical research in infectious diseases to improve the quality, relevance, recruitment, and dissemination. Further research is needed to address distinctions between acute and chronic infectious diseases in terms of patient involvement.

## Introduction

The perspective towards involving patients in clinical research has changed dramatically over the years, changing from research about patient to research with patients (1–5). Integrating the perspective of patients in clinical trials is essential to optimize clinical care. This shift was driven by the recognition that patient involvement enhances the quality, efficiency, impact, and outcomes of research (6). In addition, this shift was prompted by ethical and political considerations associated with the empowerment of patients and increased demand of accountability and transparency of public spendings (6–8). Despite this trend of increased patient involvement in clinical research, hardly any efforts have been made to improve patient involvement within the research in acute infectious diseases (9, 10).

Implementation of the perspective of patients in clinical research in infectious diseases is strived for and desired by clinicians, researchers, patient representatives, pharmaceutical industry, policy makers, and funders to improve the quality, relevance, recruitment, and dissemination of clinical research (11, 12). Most research has focused on augmenting the influence, significance, and evaluative efforts concerning the engagement of patients in clinical research. (1, 4–11, 13–28). In addition, in a review of the literature several barriers and opportunities of patient involvement in clinical research for patients and researchers were discussed (9). However, these barriers and opportunities within stakeholders beyond the researchers remain unclear. Some studies addressed the patient perspective in clinical studies in infectious diseases (9, 10, 29–31). To the best of our knowledge, a comprehensive overview of factors related to integrating the perspective of patients in the design and conduct of clinical research in infectious diseases has not been reported. In addition, several studies investigated the opinions of patients and clinical researcher about patient involvement in clinical research (10, 23, 24, 26, 27). We aim to include a wider variety of stakeholders involved in clinical research to build a comprehensive overview of opinions, ideas, and perspectives. The inclusion of various stakeholders involved in clinical research in infectious diseases is important to direct the clinical research community towards optimal integration of the patient perspective in the design and conduct of clinical research (9). The inclusion of various stakeholders involved in clinical research in infectious diseases is important to direct the clinical research community towards optimal integration of the patient perspective in the design and conduct of clinical research (9, 32).

Our qualitive study aims to explore perspectives, ideas, and opinions related to integrating the perspective of patients in the design and conduct of clinical research in infectious diseases among all stakeholders involved. In the context of this paper, we refer to patients as people who have experience with infectious disease and participation in a clinical study.

## Materials and Methods

This qualitative study was approved by the ethics committee of the University Medical Centre Utrecht (UMCU): 23U-0086. Consolidated criteria for reporting qualitative studies (COREQ) (33) are reported in supplementary table 1.

### Study population

Between March 2023 and June 2023, stakeholders involved in the field of clinical studies related to infectious disease were purposely sampled. Eligibility criteria were being a Dutch or English-speaking adult (>18 years) and experienced with infectious disease clinical studies. To ensure maximum variation, we included participants who differed by gender, age, and expertise. Sampling was scheduled to stop after a priori thematic saturation was reached (34).

### Data collection

Potential participants could contact K.A.G.J.R. to make an appointment for the interview, which was confirmed by e-mail with more information about the study aims and interview procedures. All interviews were audio-recorded and conducted via Microsoft teams (n=13). Participants received no reimbursement for their time.

Semi-structured in-depth interviews were conducted using an interview topic guide using constructs from the Consolidated Framework for Intervention Research (CFIR) (35) and feasibility framework of (36). The interviews explored perceptions and acceptability in relation to integrating the perspective of patients in clinical studies focused on infectious diseases (table 1). In addition, the implementation and practicality of the patient perspective in clinical studies were explored. The topic guide was pre-tested during three pilot interviews and subsequently revised.

**Table 1.** Interview topic guide.

| Topic | Example question or description |
| --- | --- |
| Introduction | Introduction of the interview, researcher, and participant |
| ***Intervention characteristics*** |  |
| Acceptability | Can you elaborate on the added value of the patient perspective in clinical studies? |
| Relative advantage | What are advantages of integrating the patient perspective in clinical studies?<br>What are disadvantages of integrating the patient perspective in clinical studies? |
| ***Outer setting*** |  |
| Patient needs & resources | To what extent are you aware of the needs, perspectives, and wishes of patients with regards to integrating their perspective in clinical studies? |
| Implementation | What are barriers of integrating the perspective of patients in clinical studies?<br>What are facilitators of integrating the perspective of patients in clinical studies? |
| ***Inner setting*** |  |
| Readiness for implementation | Can you elaborate on creating a patient outcome measure?<br>Can you elaborate on the process of working with patients in creating outcome measures?<br>Can you elaborate on your experience with working together with patients to integrate their perspective in clinical studies? |
| Integration | What do you need to apply the patient perspective in clinical studies? |
| ***Individual characteristics*** |  |
| Value | Can you summarize your experience with integrating the patient perspective in clinical studies? |
| Constructs and questions derived from Damschroder et al., (35, 37) and Bowen et al., (36) |  |

The interviews were conducted by K.A.G.J.R., an experienced social science researcher and I.V. a MSc medical student. The interview started with an introduction of the study and an explanation of the interview. Next, the interviewer discussed the informed consent orally with the participant and asked whether the stakeholder had any questions. Open-ended questions were used to stimulate stakeholder’s own interpretation, and stakeholders were encouraged to elaborately describe their point of view. Prompts were used to further encourage deliberation. During the interviews, notes were taken to describe nonverbal communication. Finally, the stakeholders were asked to provide background information. Interviews lasted approximately 45 minutes and were conducted in Dutch (n=12) or in English (n=1).

### Data analysis

The data analysis consisted of several stages and two cycles (38). A priori thematic saturation was evaluated after the first and second cycle. By the completion of the second cycle, a priori thematic saturation was achieved (34). The scheduled interviews were still conducted (38).

Verbatim transcription was applied to the interviews. A methodical examination of the data was carried out by K.A.G.J.R. and S.M., employing Braun and Clarke’s thematic analysis (39) to facilitate transformation of the data (40). Our analytical aim was to engage in interpretative thematic analysis, transcending mere description, and delving into the underlying explanations inherent to the data (39, 40). Independent coding of the data was undertaken by both researchers, followed by consensus-building discussions. The analytical process was fortified through a peer review with an expert in clinical research (T.t.D.) (39). Guided by the stages of qualitative analyses (39) (see table 2), recurring themes, concepts, and patterns within the data were discerned. NVivo, version 20 (41) was employed as a supportive tool for data analysis. Translations of Dutch quotations into English were meticulously executed by K.A.G.J.R. and S.M. using the forward-backward translation method. Ensuring the reliability of the data analysis, we adhered to the 15-item checklist articulated by Braun and Clarke (39). This checklist provided an additional layer of rigor to the analytical process.

**Table 2.** Stages of thematic analysis.

| Stage | Description |
| --- | --- |
| 1) Familiarization of data | K.A.G.J.R. and S.M. transcribed the data, re-read transcripts, and initial ideas were noted down. |
| 2) Generating initial codes | K.A.G.J.R. and S.M. systematically applied coding to the entire dataset using NVivo. This process involved the grouping of relevant codes in the data. |
| 3) Searching for themes | K.A.G.J.R. and S.M. engaged in a process of aligning distinct codes, orchestrating potential themes where all pertinent data were systematically coded and aggregated within these emerging themes. Following this, a deliberate exploration by K.A.G.J.R. and S.M. revealed additional patterns and repetitions within the data, facilitating a departure from the initially addressed subjects. This approach generated a thematic map. |
| 4) Reviewing themes | K.A.G.J.R. and S.M. meticulously examined coded excerpts corresponding to the identified themes, seeking out consistent patterns. Following this initial review of the coded extracts, transcripts underwent a thorough re-reading, aimed at assessing the soundness and authenticity of the identified themes. This dual review process involving the coded excerpts and transcripts ensured a rigorous evaluation of the extent to which the themes captured the essence and meaning inherent within the data. |
| 5) Defining and naming themes | K.A.G.J.R. and S.M. consistently refined each theme, ensuring that the narrative captured within the data was accurately conveyed through the refined thematic constructs. These developed themes subsequently underwent a peer review process by a clinical research expert (T.t.D.). |
| 6) Producing the report | K.A.G.J.R. and S.M. curated compelling examples – direct quotes – extracted from our data pertaining to the themes identified and research question. These illustrative quotes shaped the narrative of our comprehensive report. |
| Stages derived from the thematic analysis of Braun and Clarke (39). |  |

## Results

### Participant characteristics

Thirteen participants were scheduled and participated in the interviews. Eight of interviewed stakeholders were female and five were male. Age ranged between 30 and 59 years, with a mean age of 44.7 years old. Of the thirteen stakeholders, six were (clinical) researchers, two represented pharmaceutical companies, two were involved with policy making, and three were patient representatives.

### Synthesizing stakeholder perspectives: implications for patient involvement in clinical research about infectious diseases

Stakeholders shared their experiences and perspectives towards involvement of patients in clinical research in infectious diseases. Despite their various backgrounds and expertise, all stakeholders agreed that the patient perspective is important for the quality, relevance, recruitment, and dissemination of clinical research. They believed that patient involvement in clinical research may improve treatment guidelines. Stakeholders shared multiple facilitating and impeding factors related to patient involvement. Two main themes and several sub-themes were explored in the following sections.

#### Endeavor to transcend the existing paradigm in clinical research

Participating stakeholders were aware of recent developments related to patient involvement in clinical research in infectious diseases, such as empowerment of the public during the COVID- 19 pandemic. Within the light of these developments, researchers mentioned that patients, patient representatives, and society demand more patient involvement in research. Many stakeholders noticed a top-down demand for involvement from patients, for example, from funding parties, as illustrated by quote 1 (see table 3). Stakeholders observed that currently, funders require patient involvement in the design of studies. Clinical researchers reported the added benefit of increased involvement of patients. For instance, a stronger link between the chosen trial outcomes and the needs of patients. Despite the increased attention for patient involvement and the perceived added benefit, stakeholders observed no increase in patient involvement in clinical research in infectious diseases (table 3, quote 2).

**Table 3.**
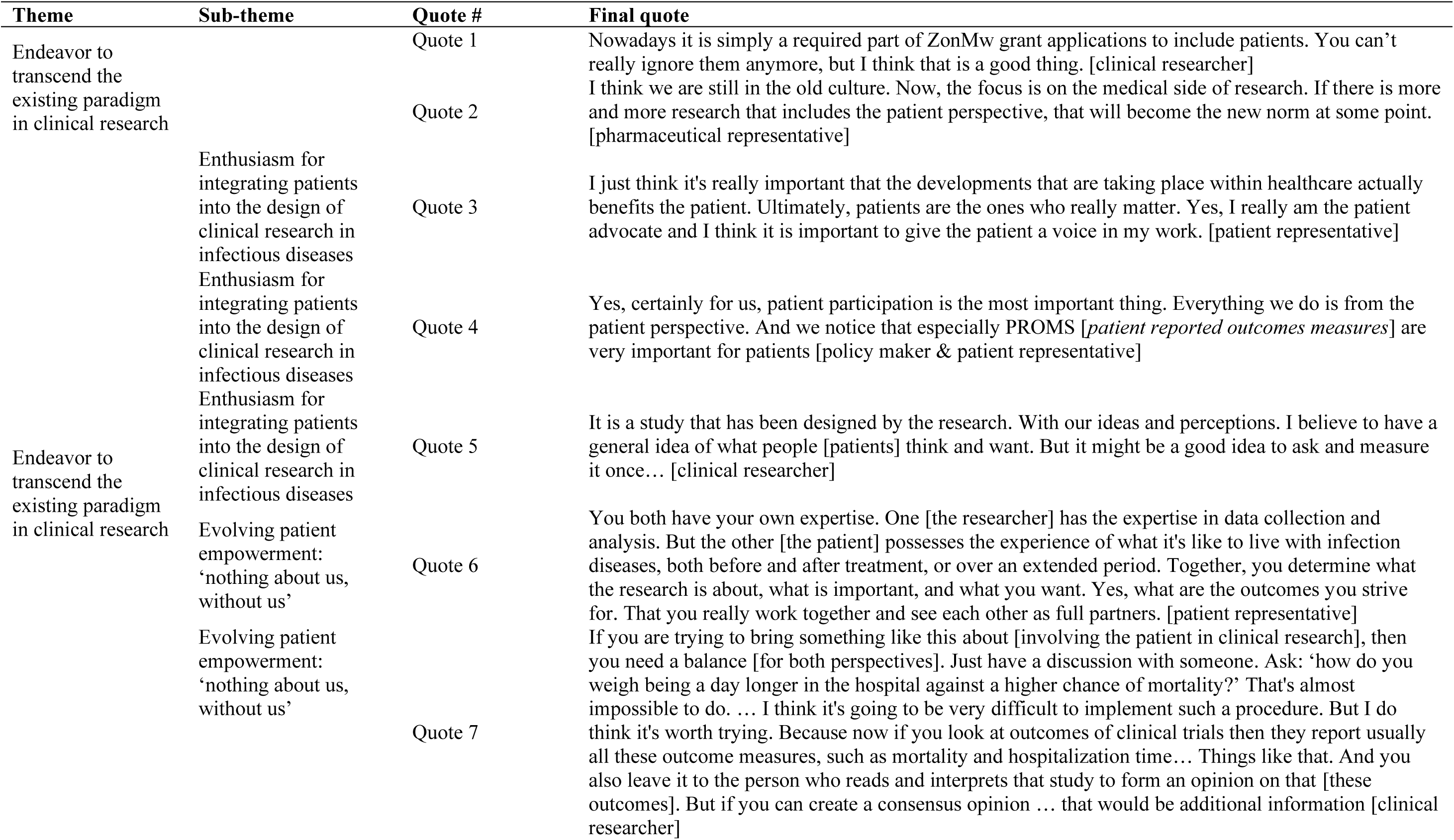
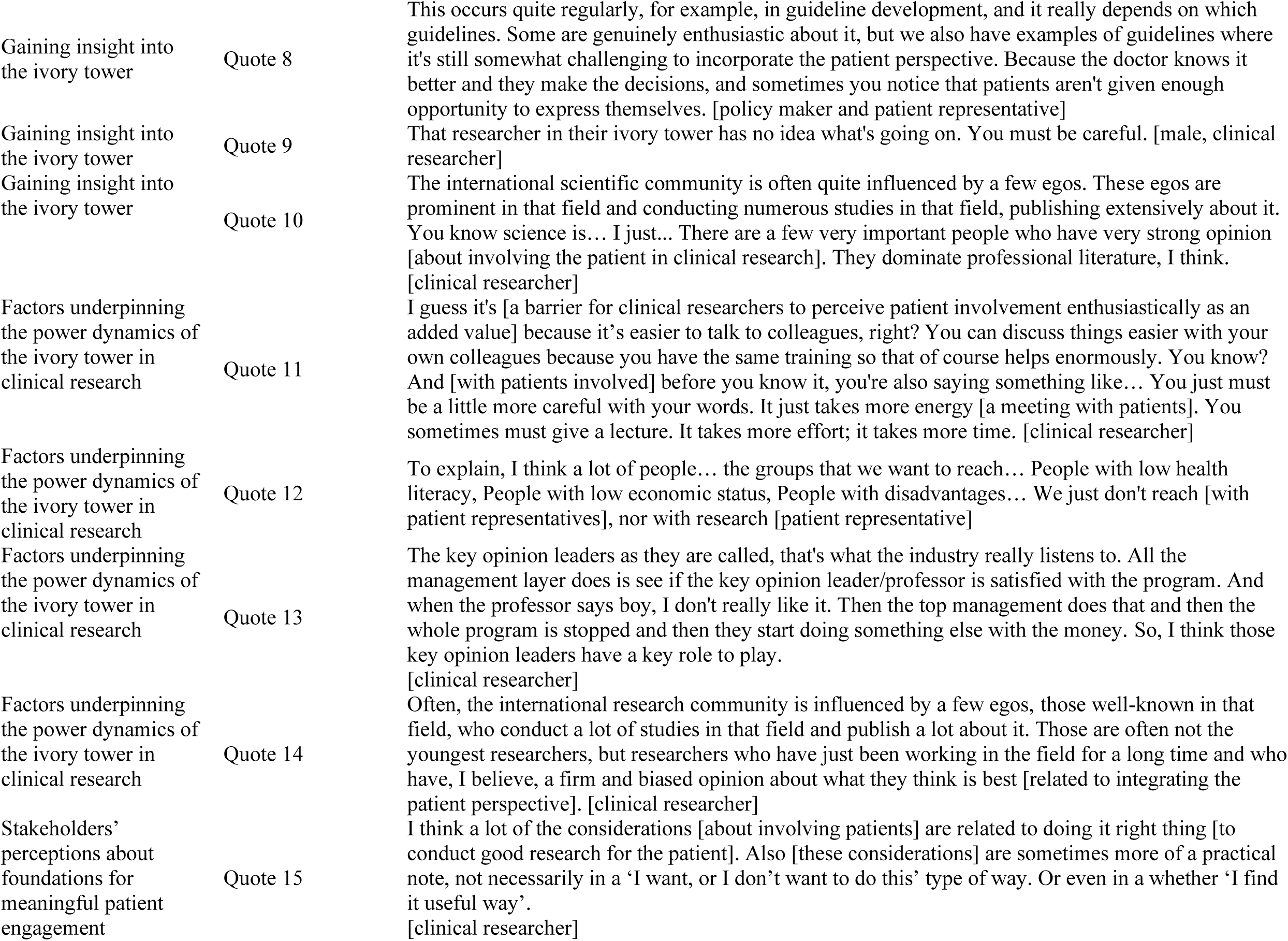
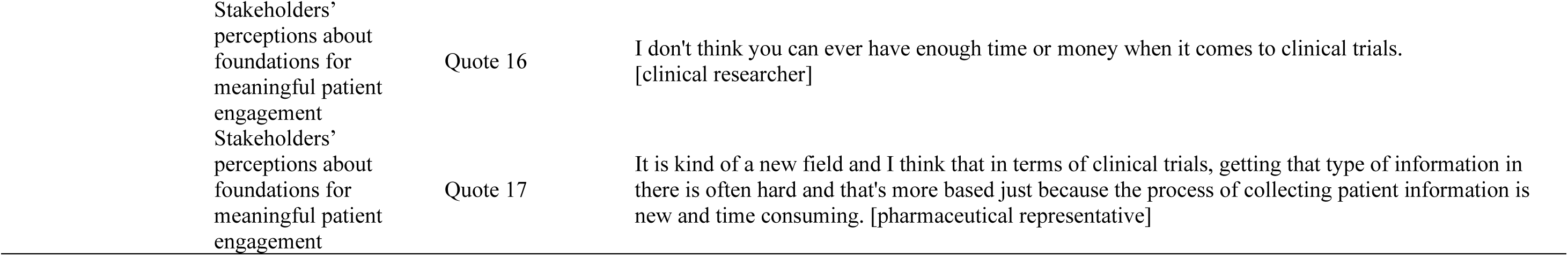
Quotes supporting endeavor to transcend the existing paradigm in clinical research.

To following sub-themes, describe and explain how stakeholders perceive the developments regarding the developments towards an increase in patient involvement in clinical research in infectious diseases.

##### Enthusiasm for integrating patients into the design of clinical research in infectious diseases

Many stakeholders shared a piqued interest in patient involvement in clinical research in infectious disease. Interest seemed to differ between stakeholders. Clinical research expressed this interest as an intrinsic motivation to ensure that healthcare is in the best interest for the patient (table 3, quote 3). Patient representatives expressed an interest to ensure that patient participation and involvement is a priority in clinical research in infectious diseases (table 3, quote 4. Some stakeholders aim to ensure that the experiences of patients are heard, and that decisions in clinical research are made based on the wishes of patients. Stakeholders emphasized that outcomes of clinical research in infectious disease are based on assumptions of clinical researchers about what patients find important (table 3, quote 5). Several stakeholders acknowledged to not explore the perception and experience of patients. For example, stakeholders explained that working with patient representatives never provides a representative overview of all affected patients, leaving room for disparity between research and end users.

##### Evolving patient empowerment: ‘nothing about us, without us’

Stakeholders explained that patients are increasingly more empowered to participate in the design and conduct of clinical research in infectious diseases. They believed that this increased empowerment was inspired by situations, such as COVID-19. Clinical researchers and patient representatives shared examples of patients striving for equality in conversations to feel taken seriously (table 3, quote 6). Striving for equality was also observed in conceptualizing the design of new clinical research Stakeholders underlined the need for important outcomes for patients, which may differ from outcomes deemed important by researchers (table 3, quote 7). This difference in perceived importance and strive towards equality was believed by stakeholders to possibly cause some unrest because it was perceived to diminish the ivory tower of research.

##### Gaining insight into the ivory tower

Stakeholders mentioned an ivory tower in research. They explained this phenomenon as a privileged position of power of clinical researchers, in which they felt – as it were – superior to patients as illustrated by quote 8 (see table 3). Often, clinical researchers make decisions about the design and conduct of clinical research without involving patients (table 3, quote 9). Stakeholders were able to observe this phenomenon in clinical research in infectious diseases and highlight key aspects of this phenomenon as explained in quote 10 (table 3) and the following sub-theme, but they were unable to determine how to resolve this phenomenon.

##### Factors underpinning the power dynamics of the ivory tower in clinical research

Stakeholders were able to identify several factors that maintain the phenomenon of an ivory tower in clinical research in infectious diseases. A persistent factor that was mentioned many times was the difference in knowledge, expertise, and language used by clinical researchers versus patients or patient representatives, as explained in quote 11 (table 3). Clinical researchers explained that a patient or patient representative is not often able to follow along with the knowledge, expertise, and language of professionals. This phenomenon was referred to as health literacy, as illustrated by quote 12 (table 3). Patient representatives underlined that, because of this gap, communication takes time and is sometimes difficult. In addition, a maintaining factor of the ivory tower phenomenon is the importance attributed to the assumptions of key researcher and clinical researchers. Stakeholders explained that for specific infectious diseases there are key opinion leaders valued by funders, such as the pharmaceutical industry (table 3, quote 13). Quote 14 explains how these key opinion leaders influence the research agenda, and thus, the conceptualization and space for involvement of patients in the design and conduct of clinical research (table 3).

##### Stakeholders’ perceptions about foundations for meaningful patient engagement

Stakeholders mentioned that several conditions are required to involve patients in clinical research in infectious diseases. Many of these conditions were labeled as practical by stakeholders, which is illustrated by quote 15 (table 3). Stakeholders often stated a lack of time and money to design and conduct research (table 3, quote 16). Next, the lack of knowledge and expertise to involve the patient perspective was emphasized by all stakeholders (table 3, quote 17). Ideally, patients, and clinical researchers would speak the same language but currently this gap was highlighted as a barrier in integrating the patient perspective in clinical research in infectious diseases. Efforts were made to bridge this gap. One patient representative explained that their organization trains patients to ease their involvement in the design and conduct of clinical research.

#### Considering the Future Landscape of Patient Involvement

Quote 18 illustrates the realization and desire of stakeholders observed to increase the involvement of patients in clinical research (table 4). Many stakeholders perceived a role for patients in the conceptualization of new clinical research in infectious disease (table 4, quote 19), but it was stressed that not every research opportunity is suited for patient involvement (table 4, quote 20). Stakeholders also noted that the role of patients cannot be the same in all types of clinical research. Barriers for patient involvement were acknowledged, and solutions to overcome these were also shared. For example, quote 21 illustrates an idea to start with patient involvement in clinical research during a medical education (table 4).

**Table 4.**
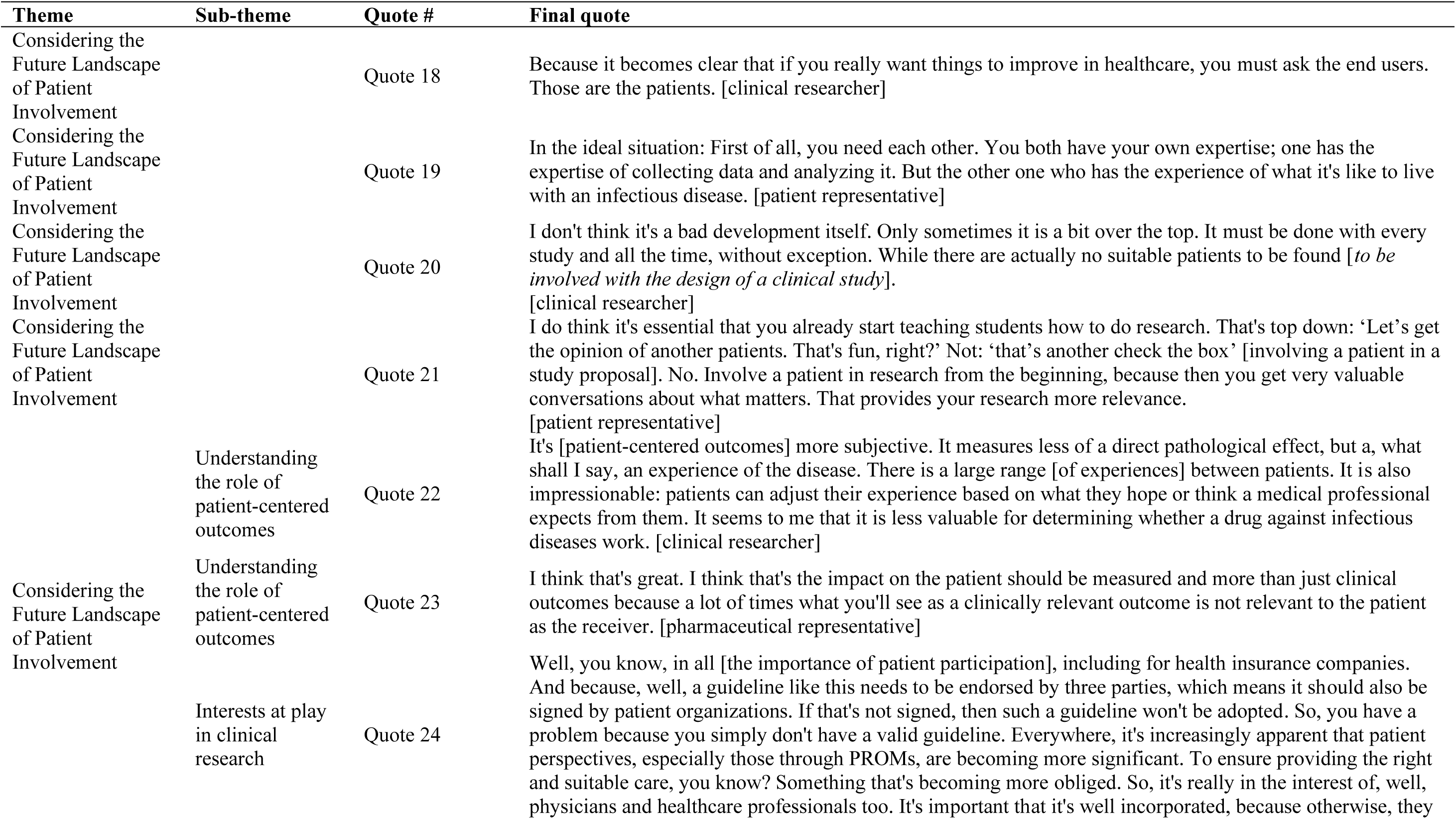
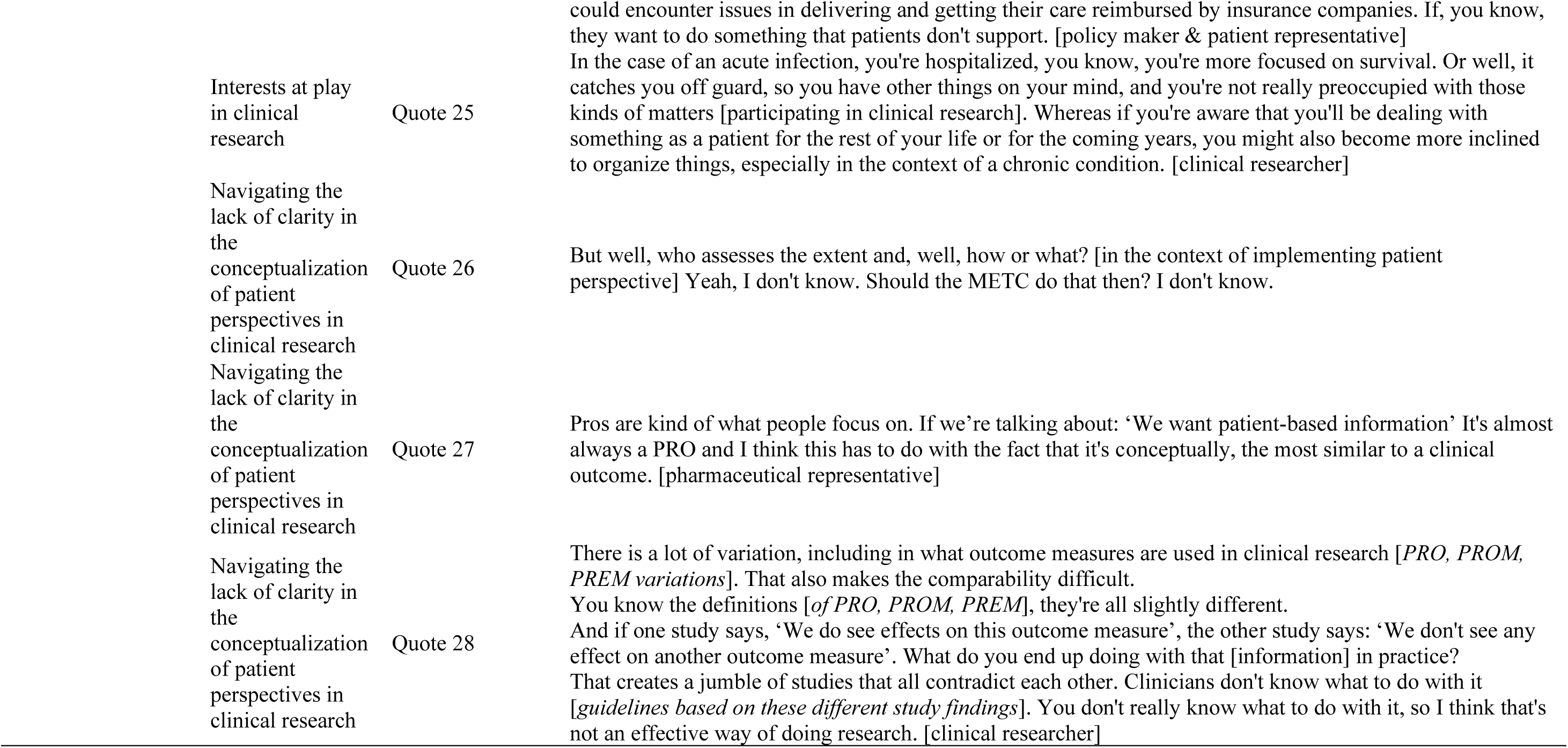
Quotes supporting considering the Future Landscape of Patient Involvement.

The following sub-themes explain in more detail facilitators and barriers perceived by stakeholders towards integrating the perspective of patients in clinical research in infectious diseases.

##### Understanding the role of patient-centered outcomes

Some stakeholders perceived patient-centered outcomes in clinical research as subjective and therefore secondary to more objective primary outcomes. This was related to the subjective experience a patient can describe compared to outcomes, such as reduced mortality. Several stakeholders perceived too much variety in the experience of patients, they believed these outcomes would not be useful in determining whether a medicine is efficacious to target an infectious disease (table 4, quote 22). However, as illustrated by quote 23, other stakeholders argued that this believe is outdated and that the impact of a new medicine to target an infectious disease should account for the relevance it offers a patient (table 4). Stakeholders acknowledged that the outcomes of clinical research result in treatment guidelines for infectious diseases. Currently, the hierarchy in determining primary and secondary outcomes is not influenced by the experience and meaning attributed by patients.

##### Interests at play in clinical research

All participating stakeholders expressed their own interests regarding the involvement of patients in the design, conduct, and impact of clinical research. Stakeholders recognized that patients, researchers, health insurance companies, and pharmaceutical companies all have their own agenda and aim to influence treatment guidelines with outcomes of clinical studies. Quote 24 explains how that the acceptance of a guideline is contingent upon obtaining approval from these different parties, where their independence on each other is apparent (table 4). Many stakeholders indicated a difference between chronic and acute infectious diseases, as shown in quote 25 (table 4). They noted that individuals with chronic infections tend to display greater interest and attribute more significance to being actively engaged in clinical research, primarily due to the chronic nature of the disease. Stakeholders believed that patients with acute infections exhibit different priorities. In addition, they observed that patients with chronic infectious diseases tend to have more knowledgeable representation, and the presence of organizations is more prevalent.

##### Navigating the lack of clarity in the conceptualization of patient perspectives in clinical research

Stakeholders agreed that the variety and lack of clarity in outcomes measured used to capture the patient perspective does not aid the implementation of these outcome measures in clinical research. Furthermore, it remains unclear to the stakeholders which entity is accountable for the implementation of the patient perspective (table 4, quote 26). Worries were expressed by stakeholders regarding the rigor of research if outcomes measures represented the patient perspective. Some researchers agreed that patient reported outcomes (PROs) resemble clinical outcomes most and thus, are usable (table 4, quote 27). Others argued that this variety and lack of clarity reduces the feasibility, measurability, and reproducibility of results. This reduced the usability for outcome measures, as it is difficult to base treatment guidelines on inconclusive results (table 4, quote 28).

## Discussion

Our thematic analyses illustrated the perceived importance of patient involvement in clinical research in infectious disease. Although barriers for integrating the patient perspective in clinical research were discussed, stakeholders underlined the importance for patient involvement for the quality, relevance, recruitment, and dissemination for clinical research in infectious Stakeholders believed money, time, expertise, clarity of conceptualization of patient involvement, and weighing perspectives of stakeholders to make designs would be needed to integrate the patient perspective in the design and conduct of clinical research in infectious diseases.

Our results demonstrate a vision stakeholders shared with regards to integrating the patient perspective in clinical research in infectious diseases. However, stakeholders were not able to identify a responsible party to ensure this vision would become a reality. A review of the literature investigated by Price, Albarqouni (9) illustrated similar findings related to unclear roles and boundaries. Many stakeholders in our study reported a lack of responsibility among involved parties, such as clinical researchers, pharmaceutical parties and regulating organizations, towards integrating the patient perspective in clinical research. A possible explanation for this lack of responsibility may be related to tensions and barriers observed (9, 13, 16, 19–21, 23, 24, 27, 42). For example, participating stakeholders seemed to perceive a tension between the wishes of patient representatives and those of clinical researchers in the choice of outcomes of clinical research. In addition, the lack of a shared language between patients and clinical researchers was perceived to hinder any form of collaboration by some. The perceived tension and barriers may reduce a feeling of responsibility and increase feelings of power among clinical researchers to integrate the perspective of patients themselves in their own clinical research.

The sub-theme “Gaining insight into the ivory tower” revealed a prevailing tendency to rely on the assumptions of key opinion leaders instead of the perspectives and experiences of patients. This was also observed in literature (6, 15, 17, 28). Many stakeholders, in particular patient representatives and clinical researchers, reflected on their respective roles in clinical research. For example, stakeholders described the tendency to rely on key opinion leaders as out-of-date from the perspective of the patient and other clinical researchers. We hypothesize that key opinion leaders could potentially serve as a facilitator in integrating the patient perspective in clinical trials (9, 35, 43). These key opinion leaders may function as champions in this process(43) and highlight external benefits, strengths, and functionality to increase patient involvement (9).

Stakeholders indicated to not being able to describe the optimal level of patient involvement in clinical research in infectious diseases. Similar findings were observed in previous research (5, 9, 16, 18). Domecq, Prutsky (16) demonstrated an absence and support of a general method for including patients in the design and conduct of clinical research. This lack of expertise for meaningful patient involvement was also observed in a mixed method study conducted among trial managers and patients in the UK (26). Our qualitative results indicate that participating stakeholders struggled with the lack of scientific methods of integrating the perspective of patients regardless of their willingness to do so. A possible explanation of the lack of the knowledge on how to integrate the perspective of patients in infectious diseases might be related to a lack of expertise in patient engagement among clinical researchers. Clinical researchers may struggle to find the best way to communicate and involve patients in the design and conduct of clinical studies (9). Thus, patients may feel left out because priorities for clinical research are fixed before they are involved (9, 10, 26).

Finally, our results demonstrate that stakeholders perceive differences in integrating the patient perspective in acute versus chronic infectious disease clinical research. Patient participation has been effectively implemented in the process of decision-making and long term management in chronic diseases (44). However, less is known about the patient involvement within the domain of acute infectious diseases. Most research in the infectious diseases has focused on chronic infections, for example, HIV. Research about HIV has made substantial effort to identify the barriers and facilitators to patient engagement in HIV research and care (45, 46). These efforts have particularly concentrated on patient-related factors and the interactions between people with HIV and physicians (11, 32, 46–49). Research focused on acute infections is limited. We hypothesize that other interests during the occurrence of an acute infection, such as the focus on survival instead of quality of life and a lack of patient representativeness, may explain the absence of patient involvement in clinical research related to acute infectious diseases. Further research is needed to explore how to integrate the patient perspective in these settings.

## Strengths & Limitations

To the best of our knowledge, this is the first study to explore the perspectives of various stakeholders about integrating the patient perspective in clinical research in infectious diseases. With triangulations with three researchers, we enhanced the credibility, transferability, dependability, and confirmability of our findings (50). There were also limitations, our qualitative results uncovered themes, issues, perspectives, and ideas of societal significance, but these findings were not intended to be generalized beyond the study domain (51). Despite variations in the stakeholders’ disciplines, expertise, and backgrounds, the qualitative nature of our study prevents us from comparing differences among stakeholders.

## Conclusions

In conclusion, our thematic analysis underlines that despite barriers, such as communication and expertise, stakeholders recognize the importance of integrating the patient perspective in clinical research in infectious diseases to improve the quality, relevance, recruitment, and dissemination. Key opinion leaders may potentially bridge these barriers and serve as champions for meaningful patient involvement. Further research is needed to address distinctions between acute and chronic infectious diseases in terms of patient involvement.

## Author declaration statement

The other authors have no competing interests to disclose.

## Author contributions

S.M.: Data collection; Data curation; Formal analysis; Project administration; Resources; Supervision; Validation; Writing – original draft; Writing - review & editing.

T.t.D: Conceptualization; Funding acquisition; Formal analysis; Supervision; Validation; Writing - review & editing.

E.S.: Conceptualization; Supervision; Validation; Writing - review & editing.

K.A.G.J.R: Conceptualization; Funding acquisition; Data collection; Data curation; Formal analysis; Investigation; Methodology; Project administration; Resources; Supervision; Validation; Writing - review & editing.

## Acknowledgements

The authors gratefully acknowledge Ilona Vriend for her input. In addition, the authors are grateful for the participants for their time.

## Funding

The authors gratefully acknowledge UMC Utrecht for funding this research.

## Ethical approval

The study was approved by the ethics committee of the University Medical Center Utrecht (UMCU): 23U-0086.

## Data availability statement

Only the authors have access to the raw data quoted in this study due to confidentiality. Any reasonable request for access to material relating to the study can be made directly to the corresponding author, who will decide on information sharing on a case-by-case basis.

## Notes

### Competing Interest Statement

The authors have declared no competing interest.

### Funding Statement

This study was funded by UMC Utrecht

### Author Declarations

Ethics committee of the University Medical Center Utrecht (U(MCU) gave ethical approval for this work (code:23U-0086)

## References

1. Chalmers I. What do I want from health research and researchers when I am a patient? Bmj. 1995;310(6990):1315-8.

2. INVOLVE. BRIEFING NOTES FOR RESEARCHERS: INVOVLING THE PUBLIC IN HS, PUBILC HEALTH AND SOCIAL CAR RESEARCH. Eastleigh: INVOLVE; 2012.

3. PCORI. Patient-centered outcomes in research 2013 [November 7, 2023:[

4. Esmail L, Moore E, Rein A. Evaluating patient and stakeholder engagement in research: moving from theory to practice. J Comp Eff Res. 2015;4(2):133–45.

5. McCarron TL, Clement F, Rasiah J, Moran C, Moffat K, Gonzalez A, et al. Patients as partners in health research: A scoping review. Health Expect. 2021;24(4):1378–90.

6. Brett J, Staniszewska S, Mockford C, Herron-Marx S, Hughes J, Tysall C, et al. Mapping the impact of patient and public involvement on health and social care research: a systematic review. Health Expect. 2014;17(5):637–50.

7. El Ansari W, Andersson E. Beyond value? Measuring the costs and benefits of public participation. Journal of Integrated Care. 2011;19(6):45–57.

8. Greene J, Hibbard JH, Sacks R, Overton V, Parrotta CD. When patient activation levels change, health outcomes and costs change, too. Health Aff (Millwood). 2015;34(3):431–7.

9. Price A, Albarqouni L, Kirkpatrick J, Clarke M, Liew SM, Roberts N, et al. Patient and public involvement in the design of clinical trials: An overview of systematic reviews. Journal of Evaluation in Clinical Practice. 2018;24(1):240–53.

10. Schilling I, Behrens H, Hugenschmidt C, Liedtke J, Schmiemann G, Gerhardus A. Patient involvement in clinical trials: motivation and expectations differ between patients and researchers involved in a trial on urinary tract infections. Research Involvement and Engagement. 2019;5(1):15.

11. Al-Shahi Salman R, Beller E, Kagan J, Hemminki E, Phillips RS, Savulescu J, et al. Increasing value and reducing waste in biomedical research regulation and management. Lancet. 2014;383(9912):176-85.

12. Chalmers I, Glasziou P. Avoidable waste in the production and reporting of research evidence. Lancet. 2009;374(9683):86-9.

13. Boote J, Baird W, Sutton A. Public involvement in the design and conduct of clinical trials: a narrative review of case examples. Trials. 2011;12(Suppl 1):A82.

14. Crocker JC, Ricci-Cabello I, Parker A, Hirst JA, Chant A, Petit-Zeman S, et al. Impact of patient and public involvement on enrolment and retention in clinical trials: systematic review and meta-analysis. Bmj. 2018;363:k4738.

15. den Houting J, Higgins J, Isaacs K, Mahony J, Pellicano E. From ivory tower to inclusion: Stakeholders’ experiences of community engagement in Australian autism research. Front Psychol. 2022;13:876990.

16. Domecq JP, Prutsky G, Elraiyah T, Wang Z, Nabhan M, Shippee N, et al. Patient engagement in research: a systematic review. BMC Health Serv Res. 2014;14:89.

17. Forsythe LP, Carman KL, Szydlowski V, Fayish L, Davidson L, Hickam DH, et al. Patient Engagement In Research: Early Findings From The Patient-Centered Outcomes Research Institute. Health Aff (Millwood). 2019;38(3):359–67.

18. Gradinger F, Britten N, Wyatt K, Froggatt K, Gibson A, Jacoby A, et al. Values associated with public involvement in health and social care research: a narrative review. Health Expect. 2015;18(5):661–75.

19. Jones EL, Williams-Yesson BA, Hackett RC, Staniszewska SH, Evans D, Francis NK. Quality of reporting on patient and public involvement within surgical research: a systematic review. Ann Surg. 2015;261(2):243–50.

20. Lander J, Hainz T, Hirschberg I, Strech D. Current practice of public involvement activities in biomedical research and innovation: a systematic qualitative review. PLoS One. 2014;9(12):e113274.

21. Oliver S, Clarke-Jones L, Rees R, Milne R, Buchanan P, Gabbay J, et al. Involving consumers in research and development agenda setting for the NHS: developing an evidence-based approach. Health Technol Assess. 2004;8(15):1–148, III-IV.

22. Oliver SR, Rees RW, Clarke-Jones L, Milne R, Oakley AR, Gabbay J, et al. A multidimensional conceptual framework for analysing public involvement in health services research. Health Expect. 2008;11(1):72–84.

23. Rhodes P, Nocon A, Booth M, Chowdrey MY, Fabian A, Lambert N, et al. A service users’ research advisory group from the perspectives of both service users and researchers. Health Soc Care Community. 2002;10(5):402–9.

24. Robbins M, Tufte J, Hsu C. Learning to “Swim” with the Experts: Experiences of Two Patient Co-Investigators for a Project Funded by the Patient-Centered Outcomes Research Institute. Perm J. 2016;20(2):85–8.

25. Robinson A. Patient and public involvement: in theory and in practice. J Laryngol Otol. 2014:1–8.

26. Selman LE, Clement C, Douglas M, Douglas K, Taylor J, Metcalfe C, et al. Patient and public involvement in randomised clinical trials: a mixed-methods study of a clinical trials unit to identify good practice, barriers and facilitators. Trials. 2021;22(1):735.

27. Stewart RJ, Caird J, Oliver K, Oliver S. Patients’ and clinicians’ research priorities. Health Expect. 2011;14(4):439–48.

28. Willyard C, Scudellari M, Nordling L. How three research groups are tearing down the ivory tower. Nature. 2018;562(7725):24-8.

29. Fernandes Agreli H, Murphy M, Creedon S, Ni Bhuachalla C, O’Brien D, Gould D, et al. Patient involvement in the implementation of infection prevention and control guidelines and associated interventions: a scoping review. BMJ Open. 2019;9(3):e025824.

30. Kemper S, Bongers M, Slok E, Schoonmade LJ, Kupper J, Timen A. Patient and public engagement in decision-making regarding infectious disease outbreak management: an integrative review. BMJ Glob Health. 2021;6(11).

31. Mbamalu O, Bonaconsa C, Nampoothiri V, Surendran S, Veepanattu P, Singh S, et al. Patient understanding of and participation in infection-related care across surgical pathways: a scoping review. Int J Infect Dis. 2021;110:123–34.

32. Edwards HA, Huang J, Jansky L, Mullins CD. What works when: mapping patient and stakeholder engagement methods along the ten-step continuum framework. J Comp Effect Res. 2021;10(12):999–1017.

33. Tong A, Sainsbury P, Craig J. Consolidated criteria for reporting qualitative research (COREQ): a 32-item checklist for interviews and focus groups. Int J Qual Health Care. 2007;19(6):349–57.

34. Saunders B, Sim J, Kingstone T, Baker S, Waterfield J, Bartlam B, et al. Saturation in qualitative research: exploring its conceptualization and operationalization. Qual Quant. 2018;52(4):1893–907.

35. Damschroder LJ, Reardon CM, Widerquist MAO, Lowery J. The updated Consolidated Framework for Implementation Research based on user feedback. Implementation Science. 2022;17(1):75.

36. Bowen DJ, Kreuter M, Spring B, Cofta-Woerpel L, Linnan L, Weiner D, et al. How we design feasibility studies. Am J Prev Med. 2009;36(5):452–7.

37. CFIR Guide. CFIR Domains for interview questions 2023 [Available from: https://cfirguide.org/guide/app/#/guide_select.

38. Guest G, Bunce A, Johnson L. How Many Interviews Are Enough? Field Methods. 2016;18(1):59–82.

39. Braun V, Clarke V. Using thematic analysis in psychology. Qualitative Research in Psychology. 2006;3(2):77–101.

40. Sandelowski M, Barroso J. Classifying the findings in qualitative studies. Qual Health Res. 2003;13(7):905–23.

41. QSR International Pty Ltd. Nvivo qualitative data analysis software. Version 20 ed 2022.

42. Tong A, Chando S, Crowe S, Manns B, Winkelmayer WC, Hemmelgarn B, et al. Research priority setting in kidney disease: a systematic review. Am J Kidney Dis. 2015;65(5):674–83.

43. Rogers EM. Diffusion of Innovations, 5th Edition: Free Press; 2003.

44. Longtin Y, Sax H, Leape LL, Sheridan SE, Donaldson L, Pittet D. Patient participation: current knowledge and applicability to patient safety. Mayo Clin Proc. 2010;85(1):53–62.

45. Koester KA, Johnson MO, Wood T, Fredericksen R, Neilands TB, Sauceda J, et al. The influence of the ’good’ patient ideal on engagement in HIV care. PLoS One. 2019;14(3):e0214636.

46. Flores D, Leblanc N, Barroso J. Enroling and retaining human immunodeficiency virus (HIV) patients in their care: A metasynthesis of qualitative studies. Int J Nurs Stud. 2016;62:126–36.

47. van Hoof M, Chinchilla K, Härmark L, Matos C, Inácio P, van Hunsel F. Factors Contributing to Best Practices for Patient Involvement in Pharmacovigilance in Europe: A Stakeholder Analysis. Drug Safety. 2022;45(10):1083–98.

48. Dawson-Rose C, Cuca YP, Webel AR, Solís Báez SS, Holzemer WL, Rivero-Méndez M, et al. Building Trust and Relationships Between Patients and Providers: An Essential Complement to Health Literacy in HIV Care. J Assoc Nurses AIDS Care. 2016;27(5):574–84.

49. SHARE - Centre for Resilience in Healthcare. A Guide to Patient and Stakeholder Involvement (PSI) in Research. 2023.

50. Nowell LS, Norris JM, White DE, Moules NJ. Thematic Analysis: Striving to Meet the Trustworthiness Criteria. International Journal of Qualitative Methods. 2017;16(1):1609406917733847.

51. Neale J, Miller P, West R. Reporting quantitative information in qualitative research: guidance for authors and reviewers. Addiction. 2014;109(2):175–6.

