## Supplementary table 1 for "Exploring factors for meaningful patient involvement in infectious disease clinical studies: A qualitative pilot study among key stakeholders"

**Title page**

Sebastiaan Moggré

Universiteitsweg 100

Utrecht, 3584 CG, The Netherlands

**Supplementary Table 1 Consolidated Criteria for Reporting Qualitative Studies(33)**

| Domain 1: Research team and reflexivity | | Pages |
|---|---|---|
| Personal Characteristics | | |
| 1. Interviewer/facilitator | Which author/s conducted the interview or focus group? K.A.G.J.R. and I.V. *conducted all the interviews.* | 5 |
| 2. Credentials | *K.A.G.J.R. PhD; I.V. MSc.* | 5 |
| 3. Occupation | *K.A.G.J.R. - Assistant Professor; I.V. – Master student.* | N/A |
| 4. Gender | *Female* | N/A |
| 5. Experience and training | *K.A.G.J.R. has extensive training and over 6 years of experience in qualitative research. I.V. was trained by K.A.G.J.R. for this project. K.A.G.J.R. observed the interviews.* | N/A |
| Relationship with participants | | |
| 6. Relationship established | Was a relationship established prior to study commencement? *No* | N/A |
| 7. Participant knowledge of the interviewer | What did the participants know about the researcher? *The researcher introduced herself and the topic at the start of the interviews.* | 5 |

| | | |
|---|---|---|
| 8. Interviewer characteristics | What characteristics were reported about the interviewer/facilitator? e.g., Bias, assumptions, reasons, and interest in the research topic?<br>*K.A.G.J.R. - "An academic social scientist." I.V. – a MSc medical student.* | 5 |
| Domain 2: study design | | |
| Theoretical framework | | |
| 9. Methodological orientation and Theory | What methodological orientation was stated to underpin the study? e.g., grounded theory, discourse analysis, ethnography, phenomenology, content analysis?<br>*A thematical analysis was performed (39, 40).* | 6 |
| Participant selection | | |
| 10. Sampling | How were participants selected? e.g., purposive, convenience, consecutive, snowball?<br><br>*Stakeholders were recruited using purposive sampling. Stakeholders were approached by the second author T.D. whether they were interested to participate in the study. In addition, K.A.G.J.R. and T.D. approached possible stakeholders through contact information published on websites.* | 5 |

| | | |
|---|---|---|
| 11. Method of approach | How were participants approached? e.g., face-to-face, telephone, mail, email?<br>*Stakeholders were approached by email to assess interest by K.A.G.J.R. and T.t.D. If stakeholders were interested in participating in the study, they were approached by phone to schedule an interview.* | 5 |
| 12. Sample size | How many participants were in the study?<br>*Thirteen interviews were conducted.* | 7 |
| 13. non-participation | How many people refused to participate or dropped out? Reasons?<br>*Of those who showed interested in the study, none dropped out.* | 7 |
| Setting | | |
| 14. Setting of data collection | Where was the data collected? e.g., home, clinic, workplace?<br>*All interviews were conducted using Microsoft Teams.* | 5 |
| 15. Presence of non-participants | Was anyone else present besides the participants and researchers?<br>*No.* | N/A |

| | | |
|---|---|---|
| 16. Description of sample | What are the important characteristics of the sample? e.g., demographic data, date?<br>*Thirteen participants were scheduled for an interview and participated in the interviews. Eight of interviewed stakeholders were female and five were male. Age ranged between 30 and 59 years, with a mean age of 44.7 years old. Of the thirteen stakeholders, six were (clinical) researchers, two represented pharmaceutical companies, two were involved with policy making, and three were patient representatives.* | 7 |
| Data collection | | |
| 17. Interview guide | Were questions, prompts, guides provided by the authors? Was it pilot tested?<br>*Semi-structured in-depth interviews were conducted using an interview topic guide.* | 5 |
| 18. Repeat interviews | Were repeat interviews carried out? If yes, how many?<br>*No.* | N/A |
| 19. Audio/visual recording | Did the research use audio or visual recording to collect the data?<br>*All interviews were audio recorded.* | 5-6 |
| 20. Field notes | Were field notes made during and/or after the interview or focus group?<br>*Notes were taken during the interviews to describe nonverbal communication.* | 5-6 |

| | | |
|---|---|---|
| 21. Duration | What was the duration of the interviews or focus group? *45 to 60 minutes.* | 5 |
| 22. Data saturation | Was data saturation discussed? *The sampling was scheduled to stop based on the principle of data saturation and exactly determined a priori.(34)* | 5-6 |
| 23. Transcripts returned | Were transcripts returned to participants for comment and/or correction? *No.* | N/A |
| Domain 3: analysis and findings | | |
| Data analysis | | |
| 24. Number of data coders | How many data coders coded the data? *Two researchers S.M. and K.A.G.J.R.* | 5-6 |
| 25. Description of the coding tree | Did authors provide a description of the coding tree? *No.* | N/A |
| 26. Derivation of themes | Were themes identified in advance or derived from the data? *No themes were identified in advance.* | N/A |
| 27. Software | What software, if applicable, was used to manage the data? *NVivo version 20.(41)* | 5-6 |

| | | |
|---|---|---|
| 28. Participant checking | Did participants provide feedback on the findings? *No.* | N/A |
| Reporting | | |
| 29. Quotations presented | Were participant quotations presented to illustrate the themes / findings? Was each quotation identified? e.g., participant number? *Yes.* | 7-17 |
| 30. Data and findings consistent | Was there consistency between the data presented and the findings? *Yes.* | 7 – 17 |
| 31. Clarity of major themes | Were major themes clearly presented in the findings? *Yes.* | 7 – 17 |
| 32. Clarity of minor themes | Is there a description of diverse cases or discussion of minor themes? *Themes and sub-themes are discussed in the results.* | 7 - 17 |
